# COVID-19 Vaccine Coverage Index: Identifying barriers to COVID-19 vaccine uptake across U.S. counties

**DOI:** 10.1101/2021.06.17.21259116

**Authors:** Anubhuti Mishra, Staci Sutermaster, Peter Smittenaar, Nicholas Stewart, Sema K. Sgaier

## Abstract

**Importance:** The United States is in a race against time to vaccinate its population to contain the COVID-19 pandemic. With limited resources, a proactive, targeted effort is needed to reach widespread community immunity.

**Objective:** Identify county-level barriers to achieving rapid COVID-19 vaccine coverage and validate the index against vaccine rollout data.

**Design:** Ecological study

**Setting:** Population-based

**Participants:** Longitudinal COVID-19 vaccination coverage data for 50 states and the District of Columbia and 3118 counties from January 12 through May 25, 2021.

**Exposure(s):** The COVID-19 Vaccine Coverage index (CVAC) ranks states and counties on barriers to coverage through 28 indicators across 5 themes: historic undervaccination, sociodemographic barriers, resource-constrained health system, healthcare accessibility barriers, and irregular care-seeking behaviors. A score of 0 indicates the lowest level of concern, whereas a score of 1 indicates the highest level of concern.

**Main Outcome(s) and Measure(s):** State-level vaccine administrations from January 12 through May 25, 2021, provided by the Centers for Disease Control and Prevention (CDC) and Our World In Data. County-level vaccine coverage as of May 25, 2021, provided by the CDC.

**Results:** As of May 25, 2021, the CVAC strongly correlated with the percentage of population fully vaccinated against COVID-19 by county (r = -0.39, p=2.2×10^−16^) and state (r=-0.77, p=4.9×10^−11^). Low-concern states and counties have fully vaccinated 26.5% [t=6.8, p=1.7×10^−7^] and 26% (t=22.0, p=2.2×10^−16^) more people, respectively, compared to their high-concern counterparts. This vaccination gap is at its highest point since the start of vaccination and continues to grow. Higher concern on each of the five themes predicts a lower rate of vaccination at the county level (all p<.001). We identify five types of counties with distinct barrier profiles.

**Conclusions and Relevance:** The CVAC measures underlying barriers to vaccination and is strongly associated with the speed of rollout. As the coverage gap between high- and low-concern regions continues to grow, the CVAC can inform a precision public health response targeted to underlying barriers.

**Key Points:** *Question:* Which U.S. counties face barriers to COVID-19 vaccine rollout, and are these communities vaccinating fewer individuals?

*Findings:* The COVID-19 Vaccine Coverage Index (CVAC) comprises five themes reflecting county-level concern for low coverage. We report rural, regional, and racial divides in exposure to these vaccination barriers. The top third of states and counties of highest concern have vaccinated 19% and 20% fewer people, respectively, compared to regions of least concern.

*Meaning:* The CVAC can help contextualize progress to widespread COVID-19 vaccine coverage, identifying underlying community-level factors that could be driving suboptimal rollout to inform precision solutions.

## Introduction

A year after SARS-CoV-2 was declared a pandemic, the U.S. started the distribution of several efficacious vaccines across the country. Rapid, widespread coverage of the COVID-19 vaccine is needed to contain the pandemic, yet there is no one solution to reach community immunity efficiently and equitably.^1^ This is far more complex than standard vaccination efforts, as the COVID-19 vaccination program brought unique logistical and acceptance challenges with the rapid deployment of a novel vaccine.^2^ Communities already operating in a limited-resource environment may struggle to rapidly vaccinate for disparate reasons.^3^ Such local delays in achieving high vaccine coverage increases the risk of the spread of new variants and prolong the path to containing the pandemic.^4,5^ With limited time and resources, decision-makers are in need of local data to proactively address these challenges rather than passively react to poor rollout performance.^6^

While vaccine hesitancy is a big hurdle to overcome in introducing a new vaccine^7–9^, there are many other reasons people who are otherwise willing, do not get vaccinated.^10^ Lessons from existing vaccination programs combined with early COVID-19 polls on vaccination intent point to myriad supply- and demand-side challenges.^11,12 2,13^ To support decision-makers in identifying communities of concern and community-specific barriers to rapid COVID-19 vaccine rollout, in February 2021, we publicly released the COVID-19 Vaccine Coverage Index (CVAC). Based on historical evidence and early COVID-19 vaccination insights, the CVAC measures community-level barriers that could hinder rapid rollout of the COVID-19 vaccine across states and counties in the U.S. Decision-makers can use the CVAC for a precision approach to proactively and cost-effectively address community-level challenges for an accelerated COVID-19 vaccine rollout. Here we describe how the index was constructed and validate it using vaccine coverage data up to May 25, 2021.

## Methods

### Index design

The index was designed using the COIN framework.^14^ We conducted a literature search up to December 17th, 2020 to identify factors associated with low vaccination coverage for existing immunization programs and factors associated with COVID-19 vaccination intention. We reviewed studies published in the last 15 years that investigated the determinants of uptake and coverage of standard vaccines in the US across child, teen, adult, and elderly populations. We also reviewed late-2020 nationally representative polls reporting COVID-19 vaccine intent stratified by e.g., demographics.^15^ From this body of evidence, we selected 28 indicators, available at the state- and county-level, with consistent evidence of an association with vaccine coverage. Indicators were grouped into five thematic domains containing a total of nine sub-themes (eTable 1). We consulted subject matter experts in national vaccine-related research and/or geospatial analysis to validate this theoretical framework and reach a final structure with expert consensus. The CVAC themes, description of indicators, and data sources are provided in supplementary materials (eMethods). CVAC data is publicly available on https://vaccine.precisionforcovid.org/.

**Table 1.**
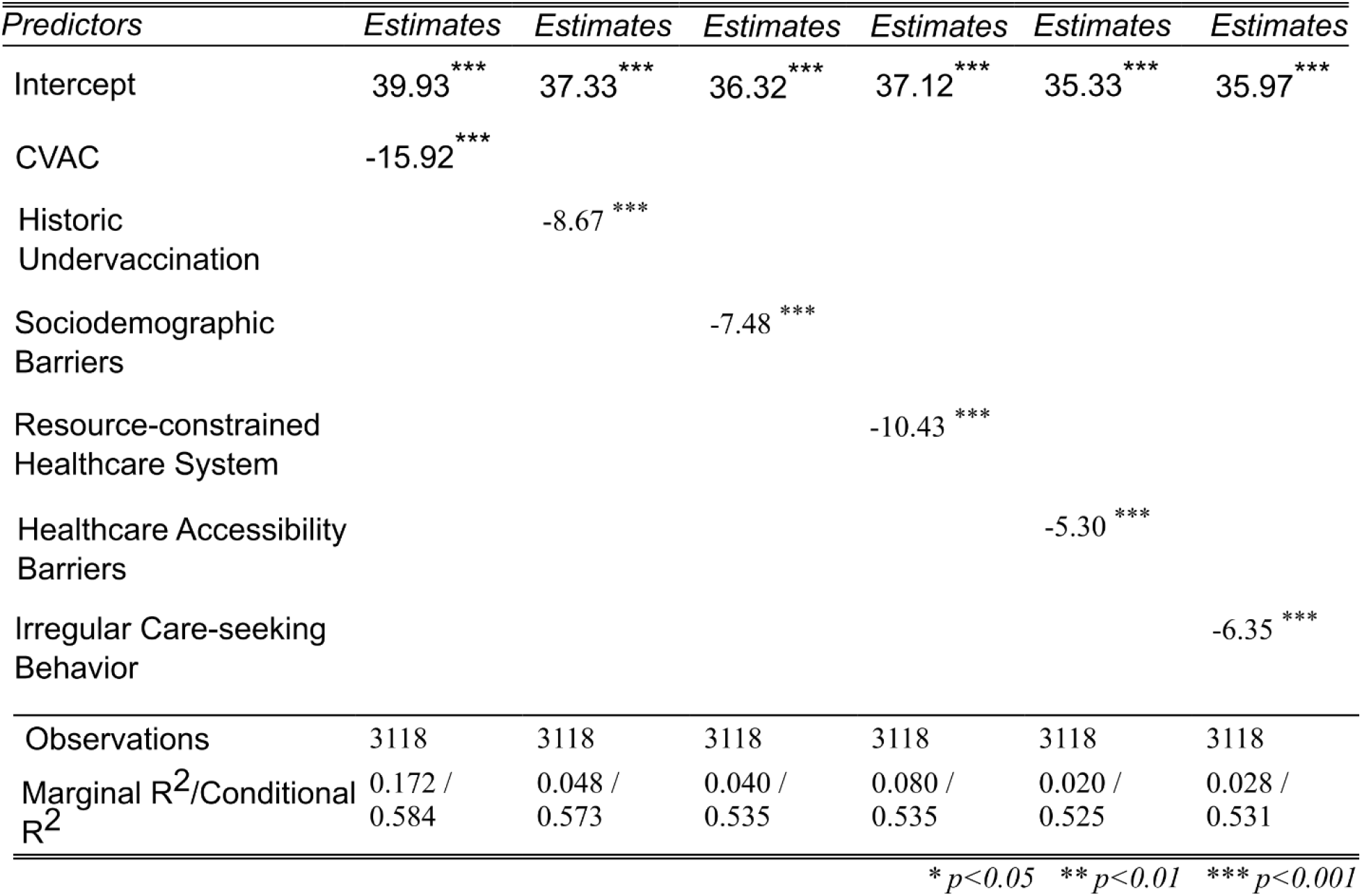
Coefficients from linear mixed-effects model with fixed-effects of the five themes and random effect of state and percentage population fully vaccinated as the dependent variable. CVAC: COVID-19 Vaccine Coverage index.

### Index computation

The CVAC was constructed at county and state levels using the same aggregation methodology as the Centers for Disease Control and Prevention’s (CDC) Social Vulnerability Index.^16^ Indicators were percentile ranked on a 0-1 scale and aggregated (i.e., summed) without further weighting into subthemes and then into themes. The themes were subsequently reranked and summed to create one composite CVAC score per state and county. State- and county-level indices were computed separately. Where indicators were only available at the state level, each county within the state was assigned the same value. Overall and theme scores were ranked on a 0-1 scale defining the relative level of concern for vaccination coverage barriers (0 = least concerning, 1 = most concerning) across geographies. Ranked scores were categorized into quintiles (very low, low, moderate, high, very high concern) and tertiles (low, moderate, high concern).

Data processing and index construction were conducted using R.^17^ With the exception of non-medical exemptions in Theme 1, less than 1% of the data was missing across all indicators. Missing data values were imputed with median values across all counties for the county-level CVAC, and across states for the state-level CVAC. For non-medical exemptions, data was missing for 70% of counties because most states do not allow for non-medical exemptions. We imputed missing values using the median exemption rate in the available data, such that this indicator only affected the CVAC in states where it could be measured.

### Statistical Analysis

Vaccination rates were calculated by CVAC tertile and plotted as a time series at state and county levels using ggplot2 in R.^18^ Spearman rank correlation coefficients were calculated using the Tidyverse package in R^19^ to assess the relationship between CVAC and vaccine rollout. CVAC scores were mapped in R^17^ to show the spatial distribution of the overall composite and barriers by theme. Geographic boundaries were defined by the 2010 Census.^20^ Using a linear mixed model, we estimated fixed-effects of individual CVAC themes and random effects of state on vaccine coverage rate across 3118 counties. We summarized the proportion of the population living in very-high CVAC counties by urbanicity, race/ethnicity, and US region using the American Community Survey 2019 population estimates.^21^ We performed a cluster analysis to group together counties with similar CVAC barrier profiles using K-medoids (partitioning around medoids).^22^ The number of clusters was selected by subjective evaluation of cluster solutions with 4 to 7 medoids. Vaccination rates per population for fully vaccinated at county level were sourced from the CDC^23^ and Texas Health and Human Services department^24^ on May 25, 2021. State-level vaccination rates were sourced from Our World in Data.^25^

## Results

### Association between vaccine coverage index and COVID-19 vaccine coverage

As of May 25, 2021, 131 million (39.9%) people had been fully vaccinated against COVID-19. However, we saw high variability in full vaccine coverage across states (mean=39.5%, SD=6.6%) and counties (mean=29.9%, SD=10.8%). Figure 1A shows the divergence in the trajectory of COVID-19 vaccination rollout of states grouped by CVAC tertiles, beginning February 1, 2021. By May 25, 2021, low CVAC (<= 0.33) states had vaccinated, on average, 26.5% (t=6.8,p=1.7×10^−7^) more people than high CVAC (>0.67) states. Examining the data for the population that had received at least one dose of the COVID-19 vaccine, we found that the gap in coverage was at its highest point (Figure 1B), starting at 5% on February 1, 2021, and rising to 22% on May 25, 2021. We found a similar relationship between CVAC and the percentage of the population fully vaccinated (r=-0.77, p=4.9×10^−11^). Looking across CVAC themes we found that states with higher barriers related to resource-constrained healthcare system (r= -0.54, p=3.6×10^−5^), healthcare accessibility (r=-0.57, p=1.1×10^−5^), and irregular care-seeking behaviors (r=-0.64, p=4.8×10^−7^) have consistently lower vaccination rates.

**Figure 1.**
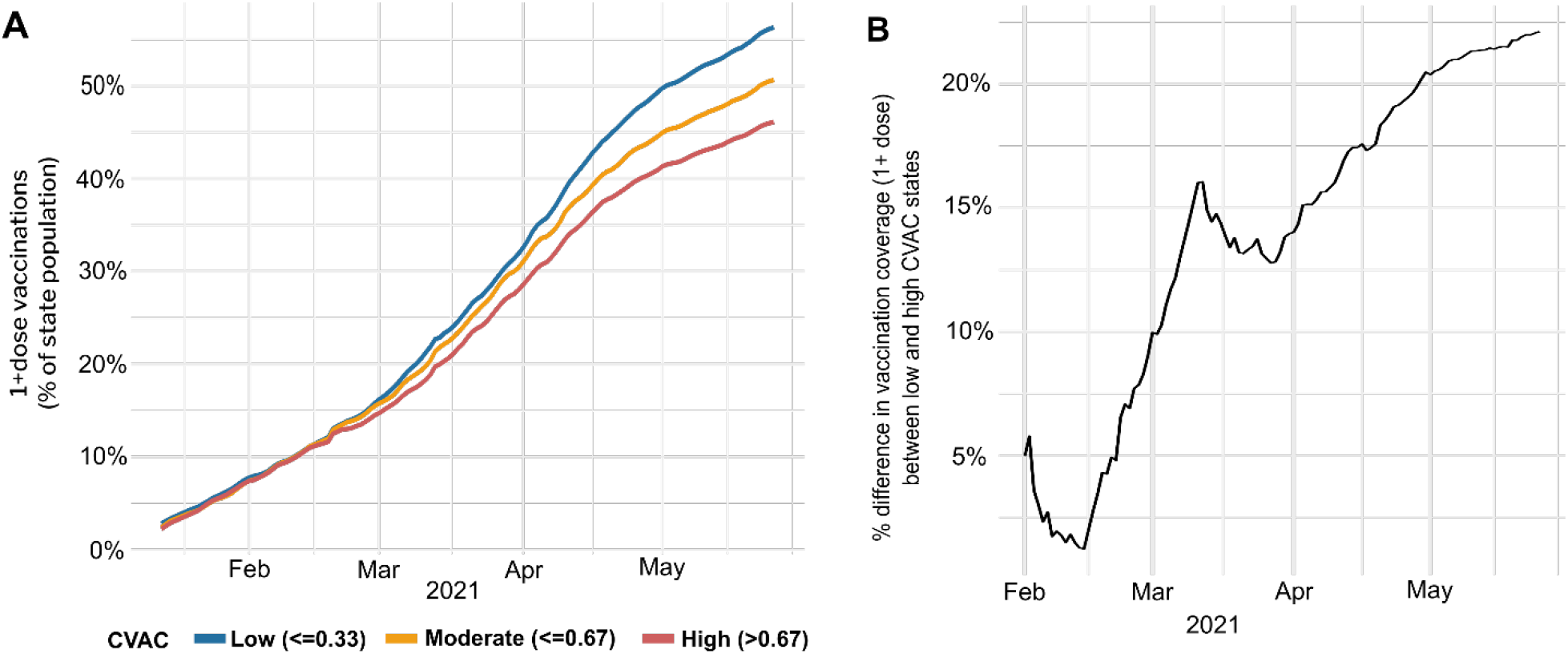
Percent of the population vaccinated over time, stratified by state-level CVAC tercile. A) Population with at least one dose per population. B) Vaccine coverage in low CVAC states, expressed relative to high-CVAC states.

### Association between CVAC and county-level vaccine *coverage*

Given the high variability in COVID-19 vaccine coverage rates across counties within states, we also examined the percentage of the population fully vaccinated for 3118 counties as of May 25, 2021^23,24^. Overall CVAC score is negatively correlated (r= -0.39, p=2.2×10^−16^) with coverage. Linear random-effects models with the CVAC and five themes as independent variables and state as random effect shows that on average, an increase from 0 (lowest concern) to 1 (highest concern) in CVAC score, is associated with a 15.9 (95% CI[-17.55, -14.29]) percentage point decline in vaccine coverage (Table 1). Similarly, we found significant associations for all five themes: Historic Undervaccination (−8.67, 95% CI[-11.14, -6.19]), Sociodemographic Barriers (−7.48, 95% [-8.53, -6.44]), Resource-constrained healthcare system (−10.43, 95% [- 11.69, -9.17]), Healthcare accessibility barriers (−5.3, 95% [-6.59,-4]), Irregular care-seeking behaviors (−6.35, 95% [-8.92,-3.78]).

### Distribution of CVAC and theme scores across the U.S

Counties with very high concern on the CVAC cluster in the South and West, where more than a third of counties in each region are of very high concern for a difficult rollout (Figure 2A). Counties in the Northeast and Midwest are on average of a lower concern than Southern and Western counties. However, when we examine concern across the five themes of CVAC, we see that regional distribution of barriers is heterogeneous with 54% of all US counties scoring “very high” on at least one of the five themes (Figure 2B).

**Figure 2.**
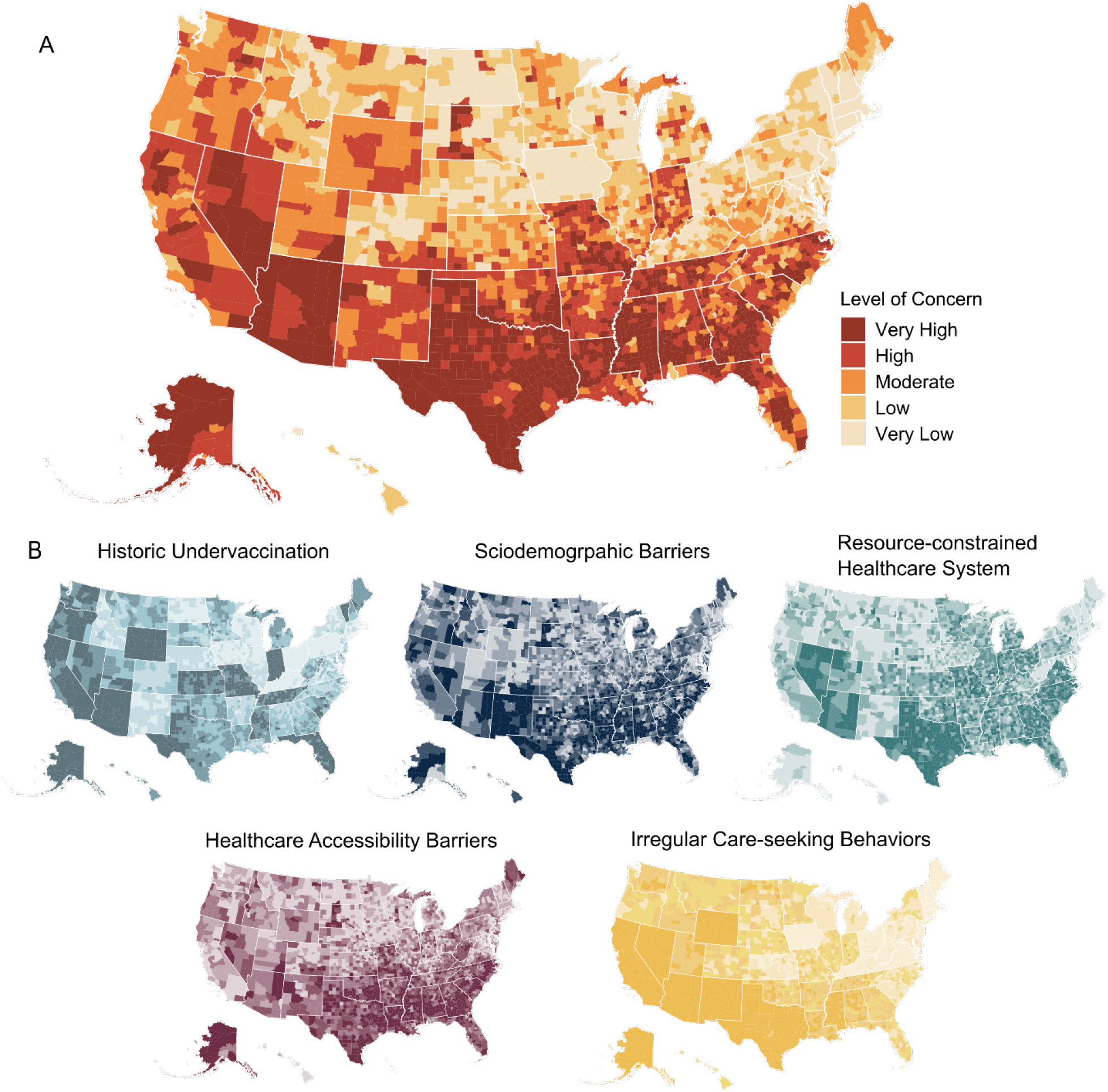
Barriers to COVID-19 vaccine coverage across 3142 US counties, A) as measured by the COVID-19 Vaccine Coverage Index (CVAC). Each quintile (very low to very high) contains 20% of US counties. B) County-level scores for each of the five themes in the CVAC. Scores are divided into quintiles from Very Low concern (lightest shade) to Very High concern (darkest shade).

### Rural, Regional, and Racial divides

There are substantial differences along urban/rural, race/ethnicity, and regional dimensions in the likelihood of living in counties facing vaccine coverage barriers (Table 2). Compared to urban communities, Americans living in rural communities are 3.7 and 2.3 times more likely to face a highly resource-constrained healthcare system and healthcare accessibility barriers, respectively. Looking across census regions, the South is particularly of high concern on healthcare accessibility barriers, where its population is more than 2 times as likely to live in counties of high concern compared with the average US population. The West is disproportionately affected by Irregular Care-seeking Behavior, with 72% of the population being of very high concern, including the majority of that region’s population—56 million—particularly in large, urban areas. American Indian and Alaska Native populations are 2.6 times more likely to live in high-concern CVAC counties than average US populations. Moreover, they consistently live in counties with high concern on all five subthemes. Similarly, Latino-Americans are twice as likely to live in communities with high-concern CVAC scores and irregular care-seeking behavior.

**Table 2.** Percentage of population living in Very High-scoring counties (top quintile) stratified by race/ethnicity, urban/rural, and U.S. region. Red color indicates a greater percentage living in Very High county, relative to the US average in the same column. Clusters of Communities Facing Similar Barriers to Vaccine Coverage.

|  | Subgroups | CVAC | Historic Under-vaccination | Socio-demographic Barriers | Resource constrained Healthcare System | Healthcare Accessibility Barriers | Irregular Care-Seeking Behavior |
| --- | --- | --- | --- | --- | --- | --- | --- |
|  | US Average | 10% | 23% | 6% | 10% | 12% | 29% |
| Urbanicity | Urban | 8% | 24% | 4% | 7% | 10% | 31% |
|  | Rural | 22% | 17% | 23% | 26% | 23% | 17% |
| Region | Midwest | 1% | 14% | 1% | 5% | 3% | 2% |
|  | Northeast | 0% | 1% | 6% | 0% | 4% | 0% |
|  | South | 20% | 20% | 11% | 19% | 27% | 30% |
|  | West | 8% | 52% | 3% | 5% | 1% | 72% |
| Race/<br>Ethnicity | White | 7% | 20% | 5% | 9% | 8% | 21% |
|  | American-Indian/<br>Alaska-Native | 26% | 30% | 28% | 14% | 28% | 45% |
|  | Asian | 4% | 24% | 2% | 4% | 5% | 44% |
|  | Black | 12% | 17% | 11% | 9% | 23% | 20% |
|  | Hispanic / Latino | 20% | 36% | 9% | 12% | 17% | 54% |
|  | Native-Hawaiian /<br>Pacific-Islander | 7% | 27% | 2% | 7% | 4% | 44% |

We performed K-medoids clustering with the five theme scores as input variables to group counties by barrier profile. The primary aim of this analysis was to simplify policy-making at the national level by reducing 3142 counties to five types with similar barriers to vaccine uptake. We identified five clusters of counties facing similar challenges to the COVID-19 vaccine rollout (Figure 3A). The five clusters were labeled based on a profile of CVAC barriers: ‘Undervaccinated with sociodemographic and accessibility barriers’, ‘Uniformly low barriers’, ‘Multi-dimensional barriers’, ‘Undervaccinated with accessibility and irregular care-seeking barriers’, and ‘Sociodemographic and healthcare access barriers’. eTable2 documents differences across the clusters based on eight profiling variables like urban-rural spread, racial and ethnic population, and vulnerability. Vaccination rollout, as of May 25, 2021, varied significantly by cluster (F=114.3, p=2×10^−16^), and was the slowest in clusters with high barriers: the largest difference was observed between the “Multi-dimensional barriers” (20% fully vaccinated) and “Uniformly low barriers” (30% fully vaccinated) counties (t=21.2, p=2.2×10^−16^). The clusters show some level of geographic clustering, e.g. “undervaccinated with accessibility and irregular care-seeking barriers” and “multi-dimensional barriers” clusters do not contain any counties from the Northeast (Figure 3B).

**Figure 3.**
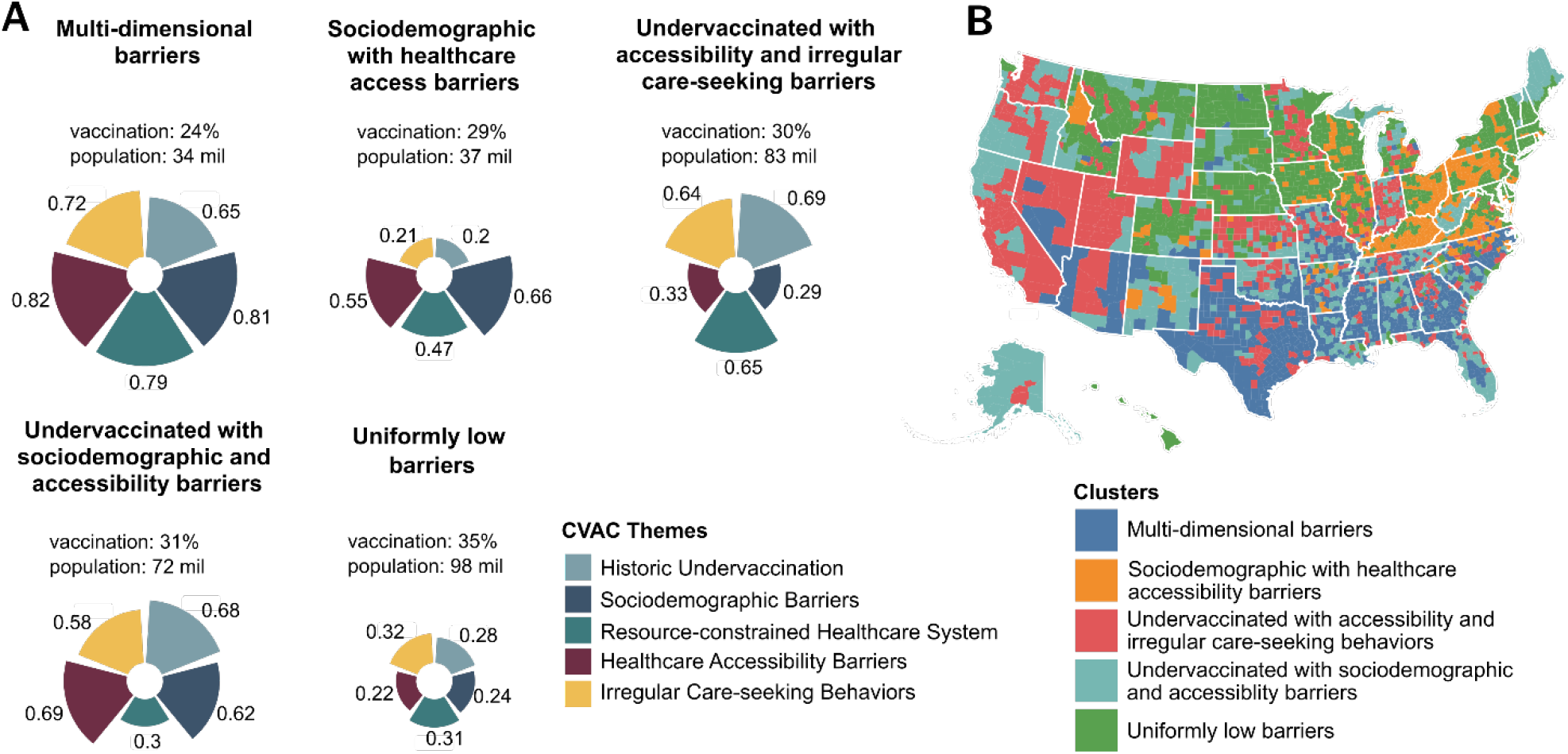
CVAC clusters with similar barriers. A) mean CVAC score by theme, % of the population fully vaccinated as of May 25, 2021, and total population. B) Geospatial distribution of CVAC clusters by county.

## Discussion

The COVID-19 Vaccine Coverage Index (CVAC) is a comprehensive index capturing community-level barriers to COVID-19 vaccine uptake in the U.S. Counties and states with high CVAC score i.e. high barriers, have vaccinated 21% fewer people than counties and states with low CVAC score. This vaccination gap between states with the highest (top third) and lowest (bottom third) CVAC barriers has only widened from 5% on Feb 1 to 26% by May 25, 2021. This adds to the concerns over community-level disparities in COVID-19 vaccine rollout, particularly in rural communities^26^, minority communities^27^, and the U.S. South and West.^28^ The CDC recently published its findings that most socially vulnerable communities have vaccinated 11% fewer adults than least vulnerable counties.^29^

Many have focused on the role of vaccine hesitancy in explaining heterogeneity in vaccine uptake^8,30^, however, our findings suggest that there are additional systemic barriers that must be addressed alongside. The CVAC offers policymakers a broader picture of supply- and demand-side factors to overcome barriers within their jurisdictions. We released the CVAC in February 2021 to support the public health community in rapidly achieving high coverage in each community. These barriers, broken down into five themes - historic undervaccination, sociodemographic barriers, resource-constrained healthcare system, healthcare accessibility barriers, and irregular care-seeking behaviors - can help formulate a targeted, precision public health response.

## Limitations

The CVAC was created based on historical evidence and early COVID-19 vaccine polls, however, the delivery of a novel vaccine is sure to present new challenges beyond what is captured in this static index. For this reason, the index is most powerful when combined with local surveillance data and local knowledge. Furthermore, the index is restricted to county-level granularity, whereas sub-county (e.g. census tract or zip code) granularity will be needed to target local pockets of undervaccination.^31^ For stakeholders working in individual counties, this index provides a first indication about prevalent barriers, as well as a framework for how to think about those barriers. However, more granular data would need to be collected to address barriers at the hyperlocal level. A limitation of vaccine coverage data used in these analyses is that they do not reflect who was vaccinated, potentially penalizing counties and states that prioritize hard-to-reach vulnerable groups.^32,33^ We intend to analyze vaccine coverage data disaggregated by age, gender, and race/ethnicity to address vaccination performance along dimensions of equity rather than only volume.

## Conclusion

County-level disparities in COVID-19 vaccine rollout are detrimental to the successful containment of the pandemic. Identifying and preparing for a multitude of barriers down to the county-level is necessary to ensure a rapid and equitable rollout. To address this challenge, we created the U.S. COVID-19 Vaccine Coverage Index (CVAC). We find the index and its themes are strongly associated with the observed rollout. County barrier profiles juxtaposed with vaccine rollout and other COVID-19 specific metrics can be used to create a precision public health response, which will help us address the challenges of this unprecedented vaccine rollout campaign proactively.

## Supporting information

eMethods

## Data Availability

The COVID-19 Vaccine Coverage Index and an interactive data explorer are available at the Surgo Venture website. COVID-19 vaccine data is available from the CDC vaccine dashboard and Texas' COVID-19 vaccination dashboard

https://vaccine.precisionforcovid.org/

https://covid.cdc.gov/covid-data-tracker/#vaccinations-county-view

https://tabexternal.dshs.texas.gov/t/THD/views/COVID-19VaccineinTexasDashboard/Summary?:origin=card_share_link&:embed=y&:isGuestRedirectFromVizportal=y

## Acknowledgments

We are grateful to the following individuals and groups for feedback on earlier versions of the index and suggestions for improvements: Amanda McClelland and the Vaccine and Community Engagement teams at Resolve to Save Lives, Puneet Dewan, Regan Marsh, and the team at Partners in Health, Daniel Salmon, Prashant Yadav, and the CDC Social Vulnerability Index team (Danielle Sharpe, Barry Flanagan, Elaine Hallisey, Grete Wilt).

## Data availability

The index is available for download at https://vaccine.precisionforcovid.org/.

