## Supplementary material for "COVID-19 Vaccine Coverage Index: Identifying barriers to COVID-19 vaccine uptake across U.S. counties": eMethods

eTable 1. Thematic structure of the COVID-19 Vaccine Coverage Index (CVAC)

| **Sub-theme** | **Indicator(s)** | **Source** | **Geo** |
| --- | --- | --- | --- |
| **Theme 1: Historic Undervaccination** | | | |
| Lower Coverage & Higher Refusal Rates | Children of age 35 months receiving ≥ 1 dose MMR vaccine | 2016-2019 National Immunization Data^a^ | State |
|  | Children of age 35 months receiving ≥ 3 doses of polio vaccine |  |  |
|  | Children of age 35 months receiving ≥ 4 doses dtap vaccine |  |  |
|  | Teens age 13-17 years up-to-date on HPV vaccine |  |  |
|  | Adults receiving flu vaccine | PolicyMap^b^ | County |
|  | Medicare beneficiaries received pneumococcal vaccine | CMS^c^ |  |
|  | Nonmedical exemption rate from child school immunization | Zipfel, 2020^d^ | County |
| **Theme 2: Sociodemographic Barriers** | | | |
| Socio- Economically Disadvantaged | Racial and ethnic minority groups (Black, Hispanic, Native Hawaiian & Pacific Islander, and Native American & Alaska Native individuals, per ACS definition) | 2014-2019 U.S. Census American Community Survey (ACS) 5-year estimates^e^ | County |
|  | Proportion of individuals without a bachelor’s degree or higher |  |  |
|  | Median household income |  |  |
|  | Average unemployment rate from March to October 2020 | Bureau of Labor Statistics^f^ |  |
|  | Individuals living below poverty level | 2014-2019 U.S. Census American Community Survey (ACS) 5-year estimates^e^ |  |
| Lack of Access to Information | Households without an internet connection |  |  |
|  | Individuals without a smartphone |  |  |
|  | Limited English-speaking households |  |  |

| **Sub-theme** | **Indicator(s)** | **Source** | **Geo** |
| --- | --- | --- | --- |
| **Theme 3: Resource-constrained Health System** | | | |
| Low Healthcare System Capacity | Vaccination provider workforce per capita (Medical Doctors, Doctors of Osteopathy, Advanced Practice Registered Nurses, Physician Assistants, Pharmacists) | HRSA, Area Health Resources^g^ | County |
|  |  | Bureau of Labor Statistics^f^ |  |
|  | Infrastructure for vaccine administration per capita (Hospitals, Urgent Care Facilities*, Veterans Health Administration Medical Facilities, Federally Qualified Health Centers and look-alikes, Pharmacies) | DHS, HIFLD^h^ | County |
|  |  | HSRA^i^ |  |
|  |  | Rx Open^j^ |  |
| Weak Healthcare System | AHRQ Prevention quality indicator (PQI) overall composite | CMS^c^ | County |
|  | Health spending per capita | CMS^c^ | State |
|  | Total healthcare funding per capita (CDC COVID Funding, Public Health Emergency Preparedness (PHEP), CDC grant for Immunization and respiratory Diseases and Vaccines for children, State Public Health Funding) | CDC^kl^ |  |
|  |  | Trust for America’s Health^m^ |  |
| **Theme 4: healthcare accessibility barriers** | | | |
| Barriers due to Cost | Individuals without health insurance coverage | 2014-2019 ACS^e^ | County |
|  | Adults who reported that there was a time in the past 12 months when they needed to see a doctor but could not because of cost. | BRFSS & KFF^n^ | State |
| Barriers due to  Transportation | Households without vehicle ownership | 2014-2019 ACS^e^ | County |
|  | Transit connectivity index (TCI) | All Transit^o^ |  |
| **Theme 5: Irregular Care-seeking Behaviors** | | | |
| Lack of a Designated Medical Home | Adults who reported that they did not have a personal doctor or health care provider. | PolicyMap^b^ | County |
|  | Children without a medical home (personal doctor or nurse, usual source for care, and family- centered care) | 2017 NSCH & KFF^p^ | State |
| Lack of Routine Care Visits | Individuals without visits to doctor for routine checkup | PolicyMap^b^ | County |
|  | Children who did not have both a medical and dental preventive care visit in the past 12 months | NSCH^q^ | State |

^a^ “NIS-Child Data Tables for 2015 to Present | CDC,” n.d., https:/[/www.cdc.gov/vacc](http://www.cdc.gov/vaccines/imz-managers/nis/datasets.html)i[nes/imz-managers/nis/datasets.html.](http://www.cdc.gov/vaccines/imz-managers/nis/datasets.html) ^b^ “Estimated Percent of Adults Vaccinated for the Flu in the Past Year in 2018.” (PolicyMap, n.d.), https:/[/www.policymap.co](http://www.policymap.com/newmaps%23/)m[/newmaps#/.](http://www.policymap.com/newmaps%23/)

^c^ Centers for Medicare & Medicaid Services, “Mapping Medicate Disparities,” 2019, https://data.cms.gov/mapping- medicare-disparities.

^d^ Casey M. Zipfel et al., “The Landscape of Childhood Vaccine Exemptions in the United States,” *Scientific Data* 7, no. 1 (November 18, 2020): 401, https://doi.org/10.1038/s41597-020-00742-5.

^e^ “2014-2019 U.S. Census American Community Survey 5-Year Estimates” (U.S. Census Bureau, n.d.). ^f^ “Labor Force Statistics from the Current Population Survey” (Bureau of Labor Statistics Data, n.d.), https://data.bls.gov/pdq/SurveyOutputServlet.

^g^ Health Resources & Service Administration, Bureau of Health Workforce, and National Center for Health Workforce Analysis, “Area Health Resource Files 2019-2020,” 2020, https://data.hrsa.gov/data/download.

^h^ Homeland Infrastructure Foundation-Level Data, “Hospitals,” 2020, https://hifld- geoplatform.opendata.arcgis.com/datasets/hospitals.

^i^ “Health Center Service Delivery and Look–Alike Sites” (Health Resources & Services Administration, n.d.), https://data.hrsa.gov/data/download.

^j^ “Facilities Map” (Rx Open, n.d.), https://rxopen.org/.

^k^ “Emergency Preparedness Funding,” n.d., https:/[/www.cdc.gov/cpr/epf/index.htm;](http://www.cdc.gov/cpr/epf/index.htm%3B) “CDC COVID-19 State, Tribal, Local, and Territorial Funding Update,” *New York*, n.d., 15.

^l^ “CDC COVID-19 State, Tribal, Local, and Territorial Funding Update.”

^m^ John Auerbach et al., “TRUST FOR AMERICA’S HEALTH LEADERSHIP STAFF,” n.d., 36.

^n^ “2013-2019 BRFSS Survey Data and Documentation” (Centers for Disease Control and Prevention, August 31, 2020), https:/[/www.cdc.gov/br](http://www.cdc.gov/brfss/annual_data/annual_2019.html)f[ss/annual_data/annual_2019.html.](http://www.cdc.gov/brfss/annual_data/annual_2019.html)

^o^ “Transit Connectivity Index (TCI)” (AllTransit), https://alltransit.cnt.org/data-download/.

^p^ “Percent of Children with a Medical Home” (The Kaiser Family Foundation, March 23, 2021), https:/[/www.kff.org/other/state](http://www.kff.org/other/state-indicator/children-with-a-medical-home/)-[indicator/children-with-a-medical-home/.](http://www.kff.org/other/state-indicator/children-with-a-medical-home/)

^q^ “Child and Adolescent Health Measurement Initiative. 2018 National Survey of Children’s Health (NSCH) Data Query.” (Health Resources and Services Administration’s (HRSA) Maternal and Child Health Bureau (MCHB), n.d.), https:/[/www.childhealthdata.org/browse/survey/allstates?q=6972#.](http://www.childhealthdata.org/browse/survey/allstates?q=6972)

### eTable 2. Cluster Profiles

| **Cluster** | **Undervaccinated with accessibility and irregular care- seeking barriers** | **Uniformly low barriers** | **Multi-dimensional barriers** | **Undervaccinated with sociodemographic and accessibility barriers** | **Socio demographic with healthcare accessibility barriers** |
| --- | --- | --- | --- | --- | --- |
| Counties | 597 counties - 19% of all counties | 866 counties - 28% of all counties | 691 counties - 22% of all counties | 560 counties - 18% of all counties | 428 counties - 14% of all counties |
| Average CVAC | **Moderate** - 0.54 | **Very Low** - 0.16 | **Very High** - 0.87 | **High** - 0.62 | **Low** - 0.37 |
| CVAC theme profile | High average scores on theme1 (0.69), theme3  (0.65), theme 5  (0.64) | Very low to low average scores all themes | Very high to high scores on all themes | High average scores on 3 themes  - th1 (0.68), th2 (0.62), th4 (0.69).  Lowest barriers on th3 (0.31) of all clusters | High average Th1 sociodemographic barriers (0.66), and moderate barriers on Th3 (0.47) and Th4 (0.55) |
| Urban/rural population | **Most people live in urban areas** (91%  of 83.4 million) | **Most people live in urban areas** (90%of 98 million) | Most people live in urban areas (68% of 34 million), **far more people live in rural counties (32%)** compared to other clusters and 75% of counties are rural | **Most people live in urban areas** (91%  of 72 million) | Most people live in urban areas (79.6% of 37 million) but **relatively high rural population (20%)** (68% of  counties are rural) |
| Regional concentration | No counties in NE - Cluster 1 counties are almost uniformly distributed across Midwest (39%),  South (34%), and  West (27%) | Majority of counties in Midwest (55%), with  13% to 16%  counties in other regions | Most counties in the South (87%) and no counties in NE | Majority of counties in the South (47%), with fewer counties in Midwest (26%),  West (23%) | Majority of counties in the South (51%), followed by Midwest (31%) amd NE  (15%) |
| Racial Profile | **High Hispanic population (21%)** but majority white population (60%) | **Highest proportion of White population (72%)** | **Majority-minority cluster (56%)** with highest Hispanic population (31%) and high black pop (17%) | **Relatively high minority pop (49%)** with 2nd highest hispanic pop (23%) and high black pop (17%) | **Relatively high while pop (65%) also high black pop (18%)** |
| Average COVID-19  vaccine coverage (2 dose) | **30%** avg vaccination rate is **relatively low** ( 26% counties missing data) | **35%** avg vaccination rate is the **highest** among 5 clusters (34% counties missing data ) | **24%** avg vaccination rate is the **lowest** among 5 clusters (17% counties missing data) | **31%** avg vaccination rate is **moderate** (35%counties missing data) | **29%** avg vaccination rate is **relatively high** (27% counties missing data) |
| US CCVI  profile | **Moderate vulnerability** (US CCVI avg = 0.41) | **Low vulnerability** (US CCVI avg = 0.26) | **High vulnerability** (US CCVI avg = 0.78) | **Moderately high vulnerability** (US CCVI avg = 0.58) | **Moderately high vulnerability** (US CCVI avg = 0.57) |

**eMethods. Supplementary Methods**

**Index Design**

The COVID-19 Vaccine Coverage Index (CVAC) is a modular index that identifies which communities are at risk for low COVID-19 vaccine coverage to inform precision solutions for an accelerated, equitable rollout. The CVAC measures community-level concern for a difficult rollout by capturing underlying supply- and demand-side barriers and unique challenges to deploying a novel vaccine. Below is a brief explanation of the contents and rationale for each CVAC theme.

**Rationale**

**Historic Undervaccination**

Standard vaccine coverage rates have been correlated with the coverage of new vaccines

during previous outbreaks^1,2^, which may also be true for the COVID-19 pandemic. Evidence consistently suggests that individuals who have not received the flu vaccine are less likely to get the COVID-19 vaccine.^3–5^ This theme captures vaccine coverage rates for flu and other standard vaccines and non-medical exemption rates (refusal of vaccines for personal reasons), which are associated with lower vaccination rates.^6–10^ The selection of indicators is described below. Though historic undervaccination is not itself a barrier, this theme was included to capture the myriad of observable and unobservable factors that could drive COVID-19 vaccine coverage rates.

**Sociodemographic Barriers**

Socioeconomically disadvantaged individuals with lower income or educational attainment or facing unemployment or poverty, have had lower vaccination coverage historically and have reported lower acceptance of the COVID-19 vaccine.^3,5,11–18^ Racial minority groups, including Black, Hispanic, and Native American individuals, have also been undervaccinated historically, have reported mixed intention in COVID-19 vaccine uptake, and have faced inequities in accessing valuable pandemic resources, such as COVID- 19 testing.^3,5,11,13–15,19,20^ This theme captures these socioeconomically disadvantaged groups as well as barriers to accessing information through proxy factors, where individuals without a cell phone or internet access or those speaking a primary language other than English are likely to have difficulty in obtaining vaccination-related information, scheduling appointments, and receiving reminders.^21(p1),22^

**Resource-constrained Health System**

Vaccinating communities quickly and widely against COVID-19 is a resource-intensive exercise. Such rapid scale-up of public health services requires high capacity and strength in the underlying public health infrastructure, human resources, and funding.^23,24^ This theme captures healthcare system capacity through potential COVID-19 vaccine distribution sites and vaccination provider workforce per capita. Different facility and provider types were selected based on CDC guidance.^25^ Weak healthcare system strength is measured by poor quality of care, low healthcare system spending, and low COVID-19 and vaccine-related funding available.

**Healthcare Accessibility Barriers**

Individuals facing difficulties in accessing care are less likely to be vaccinated because the cost and transportation barriers can discourage or prevent uptake.^26^ Higher out-of-pocket costs of vaccination have been associated with lower coverage rates and perceived as barriers to standard and COVID-19 vaccination.^16,19,27,28^ Both historical coverage rates and willingness to get the COVID- 19 vaccine tends to be lower among individuals without health insurance coverage.^11,13,15,19,20,28^ Those who have previously reported delaying care or an inability to obtain care due to cost and transportation barriers have also indicated a lower likelihood to get the COVID-19 vaccine.^26^ This theme captures both of these barriers through rates of individuals without healthcare insurance, those experiencing cost-related barriers to care or lacking household vehicle ownership, or those who live in areas with limited transport connectivity.

**Irregular Care-seeking Behaviors**

Individuals with access to routine healthcare from a provider or designated location and those who seek routine care are connected to the healthcare system and are more likely to be vaccinated.^12,13,19,20^ This theme captures these care-seeking behaviors through rates of adults and children lacking a designated medical home or failing to routinely seek care.

**Selection of Historic Undervaccination Indicators**

We considered standard adult, adolescent, and child vaccine coverage indicators. Indicators were selected based on geo-precision and strength of relationship at the most geographically granular level (e.g. county) for index inclusion. Other than county-level adult flu coverage and Medicare pneumococcal coverage, several indicators were available by age and dosage at the state level from the CDC.^29^ We selected indicators that reflected the complete dosing by age per CDC recommendations^30^, several indicators specific to key populations (including healthcare providers and nursing home populations), and indicators for the same vaccine across age groupings where available. Pearson correlation coefficients were calculated using the Tidyverse package in R^31^ to assess the relationship between vaccine coverage indicators across states. In general, states with higher coverage on one metric also scored higher on others (i.e. almost the entire correlogram is positive). Coverage rates for different vaccines were correlated within the same age group, whereas coverage rates across age groups were less correlated. The most granular indicators (i.e. adult flu and Medicare pneumococcal coverage) and indicators most correlated across age-groupings were selected for index inclusion. For example, polio vaccine coverage for children aged 35 months was highly correlated with Hepatitis B coverage for children aged 35 months (0.8), so we selected the polio vaccine indicator and discarded the other.
